# An Artificial Intelligence Model for Detection of Heart Failure with Preserved Ejection Fraction: A Report from HeartShare Study

**DOI:** 10.1101/2025.11.26.25341127

**Authors:** Ibrahim Karabayir, Shekhar Singh, Tolga Hayit, Elsayed Z. Soliman, Dalane Kitzman, David Herrington, Barry A. Borloug, Robert L Davis, Sanjiv Shah, Oguz Akbilgic

## Abstract

**Background:** Heart failure with preserved ejection fraction (HFpEF) accounts for over half of all heart failure cases in the United States and remains a diagnostic challenge. Non-invasive, scalable screening tools may enable earlier recognition, timely intervention, and improved care. To evaluate the performance, reproducibility, and early detection capability of an electrocardiogram-based artificial intelligence (ECG-AI) model designed to identify HFpEF using HeartShare data and real-world ECGs from Wake Forest Baptist Health (WFBH).

**Methods:** The original ECG-AI model was developed and validated using >1 million ECGs. In this study, we examined the external validity and reproducibility over time of this ECG-AI measure in 432 participants from an NIH-funded study of clinically validated HFpEF or controls (HeartShare). Specifically, we assessed model accuracy (AUC, sensitivity, specificity, predictive values) and reproducibility across three serial ECGs. We also analyzed the potential for early (preclinical) detection of HFpEF in 59,705 real-world ECGs from 12,338 patients a large integrated healthcare system (Wake Forest Baptist Health (WFBH)).

**Results:** In HeartShare, ECG-AI achieved an AUC of 0.760 (95% CI: 0.729-0.816), with 65% sensitivity and 75% specificity for detection of HFpEF. We obtained no significantly different AUC when using only lead I ECG as an input, AUC of 0.773 (0.729-0.816). Within-patient reproducibility across three consecutive ECGs showed strong correlations (Pearson r = 0.87-0.89) and strong agreement (Cohen’s κ = 0.68-0.74). Misclassified cases showed fewer risk factors and more normal-like ECG features. In real-world WFBH data, ECG-AI detected HFpEF up to 4 years before clinical diagnosis with AUCs from 0.77 to 0.80.

**Conclusions:** 12 lead ECG-AI model demonstrates strong generalizability, reproducibility and early detection capabilities for HFpEF, supporting its potential as a scalable screening and risk stratification tool. Almost identical single lead AUC demands future investigation for remote monitoring.

**What is new?:** This study is the first to demonstrate that an ECG-AI model for HFpEF maintains strong temporal reproducibility across serial ECGs, supporting its stability as a robust non-invasive tool. We validate the model in both a rigorously phenotyped cohort and a large real-world health system and show that single-lead ECG input achieves accuracy comparable to the full 12-lead model. In addition, we show that the model can identify HFpEF years before clinical diagnosis, extending prior work by establishing ECG-AI as a reproducible, generalizable, and potentially preclinical detection tool.

**What are the clinical implications?:** The strong temporal reproducibility of the ECG-AI measure indicates that it can provide reliable longitudinal tracking of HFpEF risk, making it suitable for both clinical monitoring and remote assessment. Early detection capabilities - up to four years before diagnosis - create opportunities for proactive evaluation and earlier intervention. The comparable performance of single-lead ECGs also opens the door for scalable deployment through wearable or home-based devices, broadening access to HFpEF screening and enabling continuous risk surveillance outside traditional clinical environments.

## INTRODUCTION

Heart failure (HF) affects approximately 6.5 million Americans, a number projected to exceed 8 million by 2030 (1–4). Over half of these cases are classified as heart failure with preserved ejection fraction (HFpEF), which is associated with high morbidity and mortality (5–7). However, the heterogeneity of HFpEF, combined with the high cost and limited accessibility of recommended diagnostic modalities, contributes to underdiagnosis and delayed diagnosis—particularly when compared to heart failure with reduced ejection fraction (HFrEF) (8,9). Unlike the declining trends observed in HFrEF, hospitalizations for HFpEF are rising, leading to longer hospital stays, increased healthcare utilization, worse outcomes, and greater costs (5,10,11).

Diagnosing HFpEF remains challenging due to its nonspecific symptoms—such as dyspnea, fatigue, and exercise intolerance—which often overlap with other conditions like pulmonary disease and obesity (12). This, coupled with the lack of accurate, noninvasive, accessible diagnostic tools, results in frequent misdiagnoses and delays in treatment (8,9,13). While right-sided heart catheterization is considered the diagnostic gold standard for HFpEF, its invasive nature, need for specialized equipment and operator expertise, and higher cost limit widespread use, emphasizing the need for more practical, accessible alternatives or prescreening tools to better select patients for invasive diagnostics in patients where the diagnosis is uncertain (14–16).

The electrocardiogram (ECG) is one of the oldest and most widely used tools in cardiovascular medicine, offering a low-cost and scalable approach to assess cardiovascular health. ECG also holds potential for early, remote assessment using smart watch or other consumer-grade electronic technology. While prior studies have explored ECG-based artificial intelligence (ECG-AI) for HF detection, most have focused on detection of reduced left ventricular ejection fraction (EF), atrial fibrillation or myopathy(17,18), or on composite outcomes encompassing all HF subtypes (19–22). Few studies have specifically targeted HFpEF detection or risk prediction using ECG-AI models (23–26). Among these, common limitations include lack of validation using clinically validated and high-quality outcomes, insufficient external validation, or limited applicability to remote monitoring due to dependence on full 12-lead ECGs. Importantly, to our knowledge, no prior work has evaluated the intra-individual temporal reproducibility of ECG-AI models.

We recently developed and validated an ECG-AI model—Wake Forest’s ECG-AI algorithm for Heart Failure—that can detect both reduced LVEF and HFpEF using 12 lead (or only lead I) data from a standard 12-lead ECG extracted from several electronic health record (EHR)-based real-world datasets (27,28). In this study, we externally validate this model using data from the HeartShare Study, which includes clinically validated HFpEF phenotyping. Additionally, we evaluate the model’s within-patient reproducibility using repeat ECGs from HeartShare participants. Lastly, we assess the early detection potential of the ECG-AI model using real world data from WFBH.

## METHODS

### Wake Forest’s ECG-AI Algorithm for LVD and HFpEF Detection

We previously developed an ECG-AI model that receive 12 lead ECG (or only lead I) (29) as an input and simultaneously returns risk of rEF (LVEF<40), mEF (40≤LVEF<50) or HFpEF (29). The model used a modified ResNet architecture (23,30) with 1D convolutional layers and trained and validated using real world data from Wake Forest Baptist Health (WFBH) in Winston-Salem, NC and University of Tennessee Health Science Center (UTHSC) in Memphis, TN (29). This study only focuses on the assessment of HFpEF detection capability of the Wake Forest’s ECG-AI algorithm for Heart Failure.

### HeartShare Study Data and Generalizability and Reproducibility Assessment

HeartShare is a NIH-funded cohort study (U54HL160273) with several other associated NIH funded projects aiming to deep phenotype HFpEF by gathering data seven sites in the United States including Northwestern University (Chicago, IL), Wake Forest School of Medicine (Winston-Salem, NC), University of Pennsylvania (Philadelphia, PA), Mayo Clinic (Rochester, MN), University of California (Davis, CA), John Hopkins (Baltimore, MD), and Mass General Brigham (Boston, MA). Participants are enrolled at all seven sites and go through a comprehensive examination including ECG, echocardiogram, labs and clinical risk factors. The cohort include both HFpEF cases with clinically validated annotation as well as controls who are free from HFpEF (31).

Electrocardiogram is a standard data modality collected during enrollment using the protocols set by Epidemiological Cardiology Research Center (EPICARE), Winston-Salem, NC and used at most other large NIH-funded cohort studies. For the purpose of a better heart rate variability assessment, ECG technicians are directed to record three consecutive ECG recordings from participants.

In this study, we used the first recorded 12 lead ECGs from each participant to validate Wake Forest’s ECG-AI algorithm for HFpEF detection. We assessed the model accuracy using area under the receiver operating characteristics curve (AUC), sensitivity, specificity, negative predictive value (NPV), and positive predictive value (PPV) statistics. Since the model outputs probabilities between 0 and 1, a threshold was selected to optimize the Youden Index, enabling binary stratification for calculating classification metrics. We further evaluated the interpretability of the model in detecting HFpEF by examining the distribution patterns of traditional ECG abnormalities among correctly classified and misclassified cases. ECG abnormalities were defined in accordance The Minnesota Code Manual of Electrocardiographic Findings (32). Categorical variables were compared using the chi-square test, while continuous variables were analyzed using the Student’s t-test.

We utilized all three consecutively recorded ECGs to assess the reproducibility of the HFpEF detection model using Pearson Correlation and Kohen’s Kappa statistics.

### Wake Forest Baptist Health Data and Assessment of Early HFpEF Recognition Potential

We gathered data from WFBH that was not used for training of the previously developed Wake Forest’s ECG-AI algorithm for heart failure detection. To evaluate baseline detection performance for HFpEF, we identified cases with ECGs recorded within 6 months prior to diagnosis. HFpEF was defined consistent with our prior work: a documented diagnosis of HF in the EHR along with an echocardiogram showing preserved EF (EF > 50%) within ±30 days of the diagnosis date.

We assessed model performance by calculating AUCs across multiple prediction windows leading up to the diagnosis. Comparisons were made between AUCs derived from earlier ECGs and the baseline AUC (i.e., ECGs within 6 months before diagnosis) to evaluate how early HFpEF could be detected without a substantial drop in accuracy. For control participants, all available ECGs across all time windows were included. Statistical comparisons of AUCs were performed using DeLong’s test (33).

To enable risk stratification, the ECG-AI output was divided into quartiles based on a pre-determined cut-off value previously defined for binary classification. Mid-rank thresholds derived from this cut-off were used to define four risk groups: low, medium, high, and very high risk. These categories were then used in downstream survival analyses.

Survival analyses included Kaplan-Meier estimators for cumulative incidence and Nelson-Aalen estimators for cumulative hazard. A Cox proportional hazards model was employed to estimate the association between ECG-AI driven risk groups and time to HFpEF diagnosis. The model was also adjusted for sex (male), race (Black or African American), and age. For HFpEF cases, time-to-event was defined as the interval between ECG acquisition and the date of HFpEF diagnosis. For non-HFpEF participants, two approaches were used:

– For those without any HF diagnosis and no recorded EF < 50%, time-to-event was defined as the duration from ECG acquisition to the last follow-up date.
– For those with an EF < 50%, the time-to-event was defined as the interval between the ECG and the earliest date when EF was documented as <50%. To avoid reverse causation, any ECGs recorded after the first EF < 50% were excluded from analysis.

## RESULTS

### External Validation on HeartShare Data

HeartShare Study data included ECGs from 432 participants including 72% White, 1.4% Black or African American, 3% Asian, 39% male, with a mean±SD age of 67±13 years old. 241 (56%) of participants were HFpEF cases and 191 (44%) controls.

Without any modification, the original ECG-AI model was applied to HeartShare 12 lead ECGs and resulted in AUC of 0.760 (0.715-0.809). Using the threshold of 0.0119 (at an optimal Youden Index of 0.39), we obtained a sensitivity of 65%, specificity of 74%, PPV of 76% and NPV of 63%. However, such threshold selection is a subjective way of decision making based on predicted probabilities. Therefore, we also calculated sensitivity and specificity at two different cutoff points one maximizing sensitivity at 50% specificity level and one maximizing specificity at minimum 50% sensitivity level. The thresholds of 0.0063 yielded a sensitivity of 82% at specificity of 50% and the threshold of 0.0177 yielded a specificity of an 87% at sensitivity of 50%.

### ECG Characteristics of Misclassification Patterns

There were 141 true negatives (TN), 50 false positives (FP), 84 false negatives (FN), and 157 true positives (TP). For HFpEF cases, we compared the eight traditional ECG characteristic between TPs and FNs including ventricular rate, atrial rate, PR Interval, QRS Duration, Corrected QT, P Axis, R Axis, and T Axis. Among eight ECG characteristics considered, seven of them showed significant difference at levels of p<0.001, p<0.01, or p<0.05 between TP and FN as summarized in Table 1.

**Table 1.**
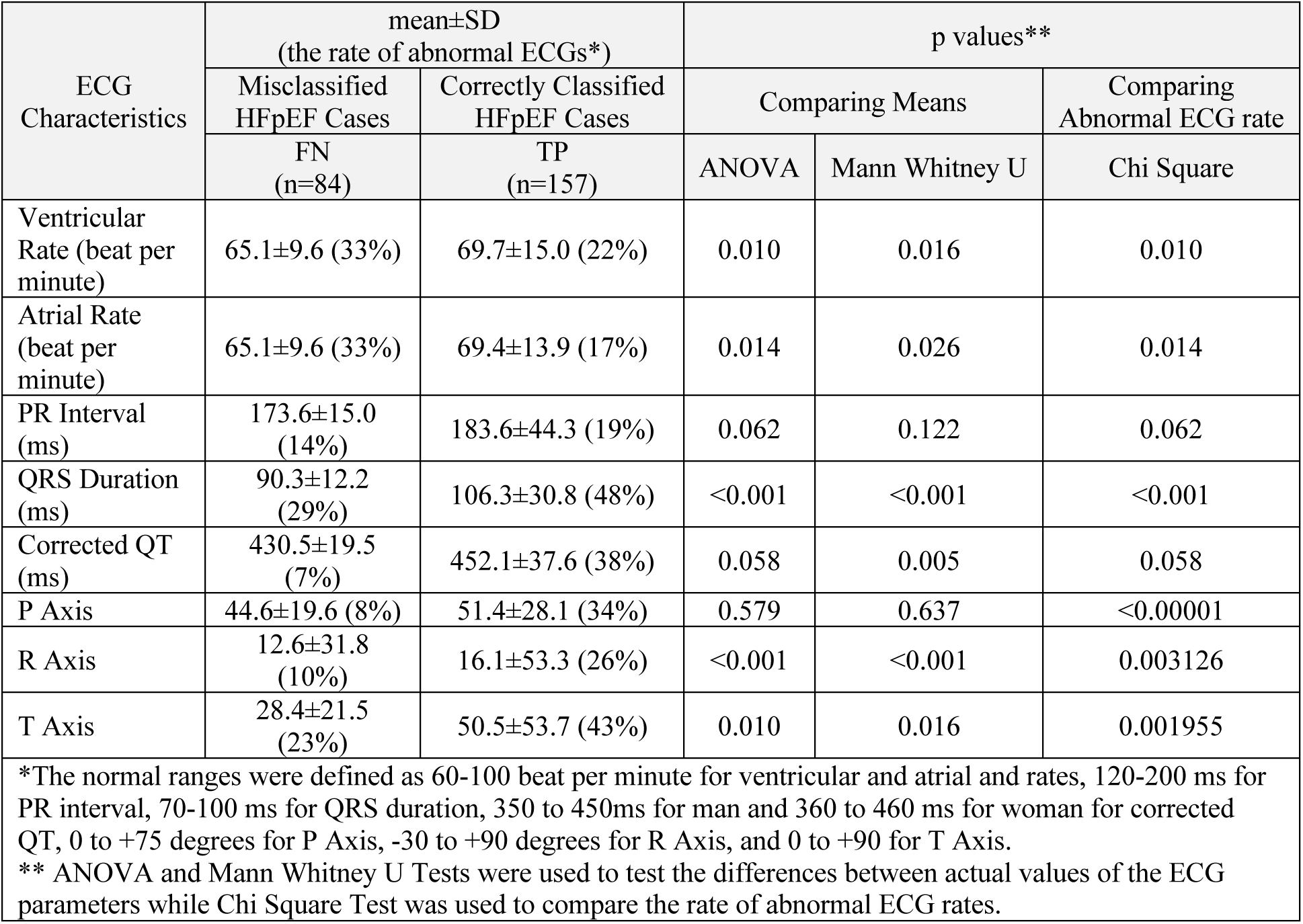
Misclassification patters by ECG characteristics.

| ECG Characteristics | mean±SD<br>(the rate of abnormal ECGs*) |  | p values** |  |  |
| --- | --- | --- | --- | --- | --- |
|  | Misclassified HFpEF Cases | Correctly Classified HFpEF Cases | Comparing Means |  | Comparing Abnormal ECG rate |
|  | FN<br>(n=84) | TP<br>(n=157) | ANOVA | Mann Whitney U | Chi Square |
| Ventricular Rate (beat per minute) | 65.1±9.6 (33%) | 69.7±15.0 (22%) | 0.010 | 0.016 | 0.010 |
| Atrial Rate (beat per minute) | 65.1±9.6 (33%) | 69.4±13.9 (17%) | 0.014 | 0.026 | 0.014 |
| PR Interval (ms) | 173.6±15.0 (14%) | 183.6±44.3 (19%) | 0.062 | 0.122 | 0.062 |
| QRS Duration (ms) | 90.3±12.2 (29%) | 106.3±30.8 (48%) | <0.001 | <0.001 | <0.001 |
| Corrected QT (ms) | 430.5±19.5 (7%) | 452.1±37.6 (38%) | 0.058 | 0.005 | 0.058 |
| P Axis | 44.6±19.6 (8%) | 51.4±28.1 (34%) | 0.579 | 0.637 | <0.00001 |
| R Axis | 12.6±31.8 (10%) | 16.1±53.3 (26%) | <0.001 | <0.001 | 0.003126 |
| T Axis | 28.4±21.5 (23%) | 50.5±53.7 (43%) | 0.010 | 0.016 | 0.001955 |
| *The normal ranges were defined as 60-100 beat per minute for ventricular and atrial and rates, 120-200 ms for PR interval, 70-100 ms for QRS duration, 350 to 450ms for man and 360 to 460 ms for woman for corrected QT, 0 to +75 degrees for P Axis, -30 to +90 degrees for R Axis, and 0 to +90 for T Axis.<br>** ANOVA and Mann Whitney U Tests were used to test the differences between actual values of the ECG parameters while Chi Square Test was used to compare the rate of abnormal ECG rates. |  |  |  |  |  |

As summarized in Table 1, several ECG-derived features—including ventricular rate, atrial rate, QRS duration, corrected QT interval (QTc), and T-wave axis—differ significantly between HFpEF cases that were correctly classified versus misclassified by the ECG-AI model.

### HFpEF Risk Factor Characteristics of Misclassification Patterns

In parallel to the ECG based misclassification analysis, we also carried out a complementary analysis by focusing on traditional HFpEF risk factors (Table 2).

**Table 2.** Misclassification Patterns by HFpEF Risk Factors.

| Condition | Description | Misclassified<br>HFpEF Cases (FN) | Correctly Classified<br>HFpEF Cases (TP) | Chi-Square (p<br>value) |
| --- | --- | --- | --- | --- |
| Heavy | BMI>30 kg/m<sup>2</sup> | 69.1% | 74.3% | 0.509 |
| Hypertension | 2 or more medication | 63.2% | 77.1% | 0.027 |
| Atrial Fibrillation | Paroxysmal or persistent | 25.0% | 54.9% | <0.001 |
| Pulmonary<br>Hypertension | Doppler echocardiogram<br>estimated Pulmonary Artery<br>Systolic Pressure >35mmHg | 25.0% | 36.6% | 0.063 |
| Elder | Age>60 years | 86.8% | 94.9% | 0.053 |
| Filling Pressure | Doppler echocardiographic<br>E/e'>9 | 52.9% | 71.5% | 0.007 |

Table 2 suggest that correctly classified HFpEF cases by ECG-AI have significantly more hypertension (p<0.05), atrial fibrillation (p<0.001) and more increases filling pressure defined as E/e’>9 (p<0.01).

### Reproducibility of ECG-AI in HFpEF Detection

The reproducibility of ECG-AI for HFpEF was assessed using Pearson Correlation and Cohen’s Kappa statistics (Table 3). Three ECG-AI based HFpEF probabilities were calculated for each participants using their consequentially recorded three ECGs. Correlation analysis suggested strong correlation (all p values <0.001) between HFpEF risk calculated from three ECGs. (Figure 1). The Pearson Correlations of 0.87, 0.89, and 0.87 were obtained for HFpEF probabilities between first and second ECGs, first and third ECGs, and second and third ECGs, respectively, all statistically significant (p<0.001). We also calculated the predicted label (at HFpEF risk or not at HFpEF risk) using the threshold of 0.0096 that was reported above. We obtained the Cohen’s Kappa statistics of 0.74, 0.68, and 0.71 between predicted HFpEF risk classes (low or high) between first and second ECG, first and third ECG, and second and third ECGs, respectively. All Kappa statistics suggested robust agreement between HFpEF risk classification from three sequential ECGs.

**Figure 1.**
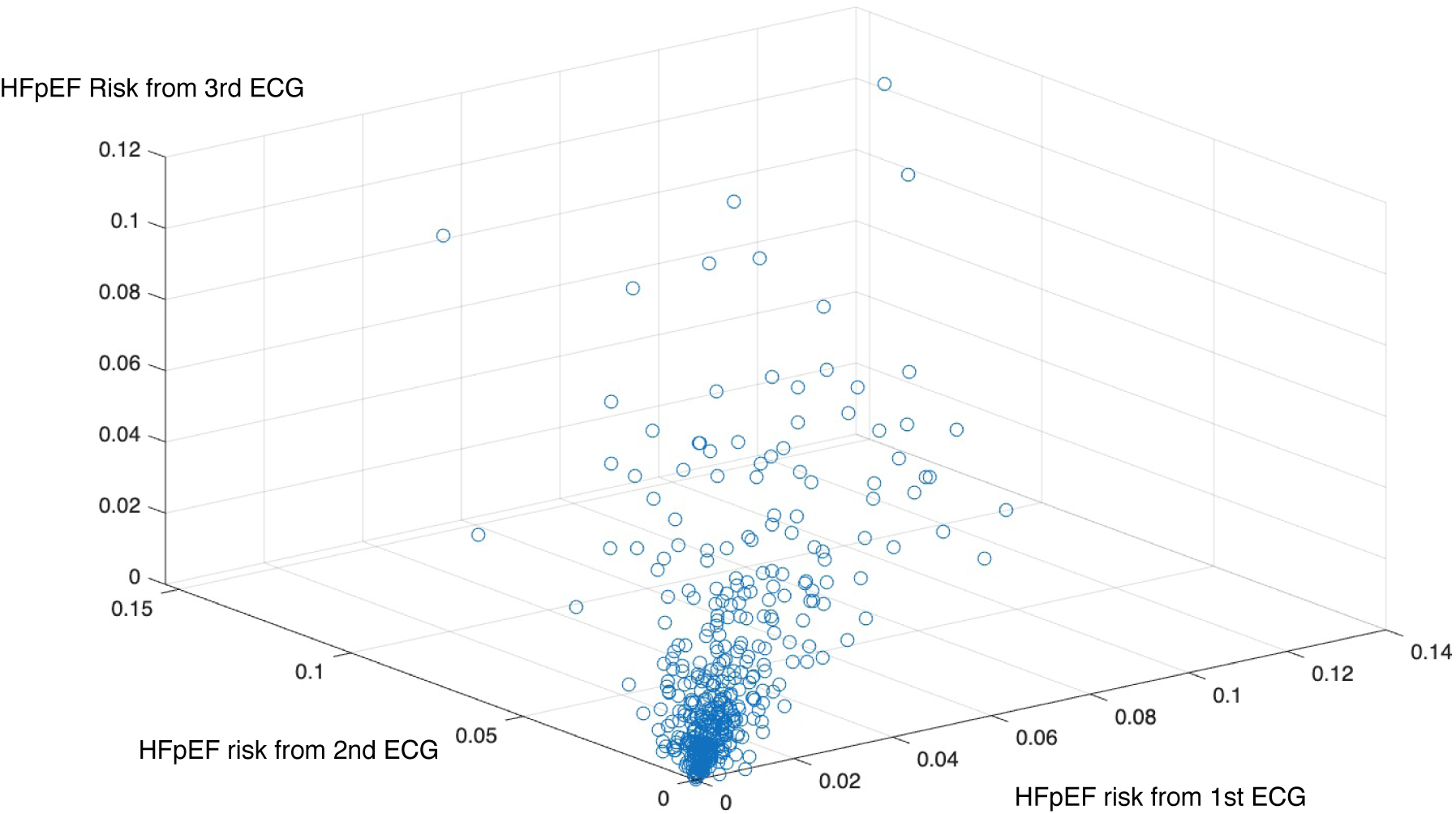
Scatter Plot of HFpEF Predictions for sequentially recorded three ECGs.

**Table 3.** Concordance of ECG-AI based HFpEF Assessment across sequentially collected ECGs.

| Concordance Method | ECGs in Comparison |  |  | Interpretation |
| --- | --- | --- | --- | --- |
|  | 1<sup>st</sup> and 2<sup>nd</sup> ECGs | 1<sup>st</sup> and 3<sup>rd</sup> ECGs | 2<sup>nd</sup> and 3<sup>rd</sup> ECGs |  |
| Pearson Correlation of Predicted Risk | 0.87 | 0.85 | 0.82 | Strong Correlations |
| Cohen's Kappa of Predicted HFpEF Status | 0.74 | 0.68 | 0.71 | Substantial Agreement |

### Comparison of 12 lead and lead I models

Despite this study focuses on HFpEF detection model utilizing 12 lead 10 second ECG, the original ECG-AI model has two separate versions, one designed for 12 lead input and one for single lead (lead I). When we tested the single lead version of the ECG-AI model on the same HeartShare data, we obtained an almost identical AUC of 0.773 (0.729-0.816). This AUC obtained using was not statistically different than the AUC of 0.760 (0.715-0.809) based on DeLong Test (p=0.681).

### Assessment of Early HFpEF Recognition Potential of ECG-AI

The analytical cohort comprised 59,705 ECGs from 12,338 patients at Wake Forest Baptist (51% female, 20% Black, mean age 58 ± 15 years) (Table 4). Of these, 922 ECGs (1.5%) were from 305 patients who were diagnosed with HFpEF after the index ECG. As a baseline HFpEF detection accuracy, we used ECGs obtained within six months of the clinical diagnosis. In this group the ECG-AI model achieved an AUC of 0.80 (95% CI: 0.78–0.83) for HFpEF detection. Next, we excluded the cases of HFpEF occurring within 6 months following the index ECG and calculated AUCs over longer intervals as summarized in Figure 2. We note that the control ECGs were the same for AUC calculation at each time interval while the case ECGs differed. AUCs were 0.80 (95% CI: 0.77–0.83) at 6 months–1 year, 0.79 (95% CI: 0.76–0.81) at 6 months–2 years, 0.77 (95% CI: 0.75–0.79) at 6 months–3 years, and 0.77 (95% CI: 0.75–0.78) at 6 months–4 years before diagnosis, suggesting that the model could detect HFpEF up to four years before clinical diagnosis without significant loss of accuracy (DeLong’s test p > 0.01). AUCs for fixed time windows (6 months to 1 year, 1 year to 2 years, etc.) were depicted in Figure 2.

**Figure 2.**
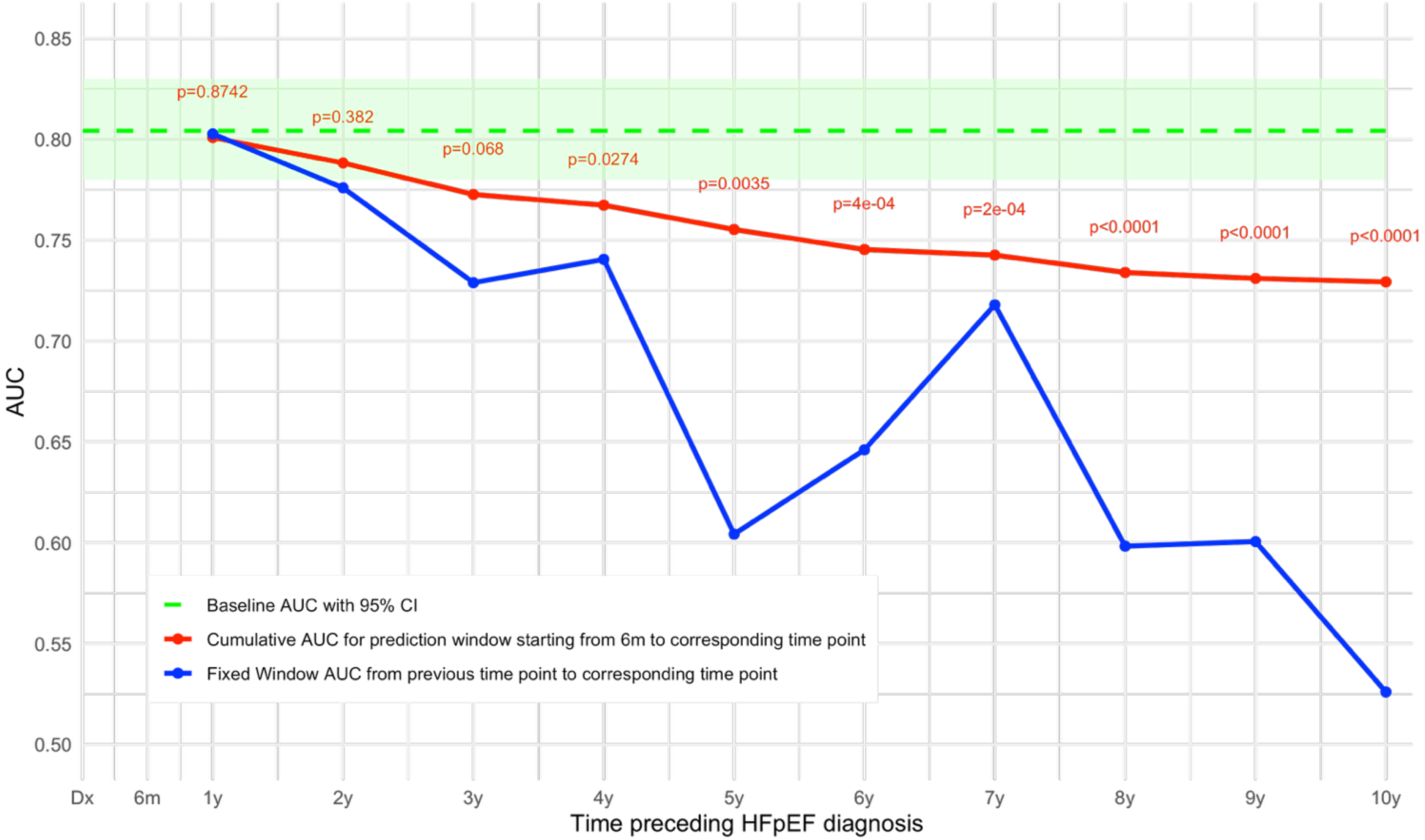
ECG-AI’s Accuracy when using ECGs at different time points.

**Table 4.** Baseline characteristics of the study cohorts based on unique ECG-level observations.

|  | Variable | HeartShare |  |  | WFBH |  |  |
| --- | --- | --- | --- | --- | --- | --- | --- |
|  |  | Control | Case | p-value | Control | Case | p-value |
| Demographics | Age | 61.0 ± 14.0 | 71.5 ± 10.0 | <0.001 | 58.3 ± 15.3 | 65.6 ± 14.5 | <0.001 |
|  | Male | 76 (40.9%) | 85 (37.3%) | 0.521 | 28841 (49.1%) | 390 (42.3%) | <0.001 |
|  | Black or African American | 28 (14.7%) | 38 (15.8%) | 0.855 | 13508 (23.0%) | 222 (24.1%) | 0.455 |
| Comorbidities | Chronic kidney (renal) disease | 5 (2.6%) | 39 (16.2%) | <0.001 | 2066 (3.5%) | 92 (10.0%) | <0.001 |
|  | Diabetes | 25 (13.1%) | 53 (22.0%) | 0.024 | 6929 (11.8%) | 315 (34.2%) | <0.001 |
|  | Coronary artery disease | 18 (9.4%) | 54 (22.4%) | 0.001 | 4059 (6.9%) | 206 (22.3%) | <0.001 |
|  | Myocardial infarction | 4 (2.1%) | 25 (10.4%) | 0.001 | 5011 (8.5%) | 310 (33.6%) | <0.001 |

The median follow-up duration among participants without HFpEF was 8.9 years (IQR: 4.7–14.0), whereas the median time to event among those who developed HFpEF was 1.7 years (IQR: 0.7–3.9). As illustrated in Figures 3, which display the Kaplan-Meier and Nelson-Aalen estimators respectively, the ECG-AI–derived risk groups show clear separation. Participants in the highest quartile of ECG-AI output (“Very High Risk”) experienced the greatest risk of developing incident HFpEF, while those in the lowest quartile (“Low Risk”) exhibited the greatest HFpEF-free survival.

**Figure 3.**
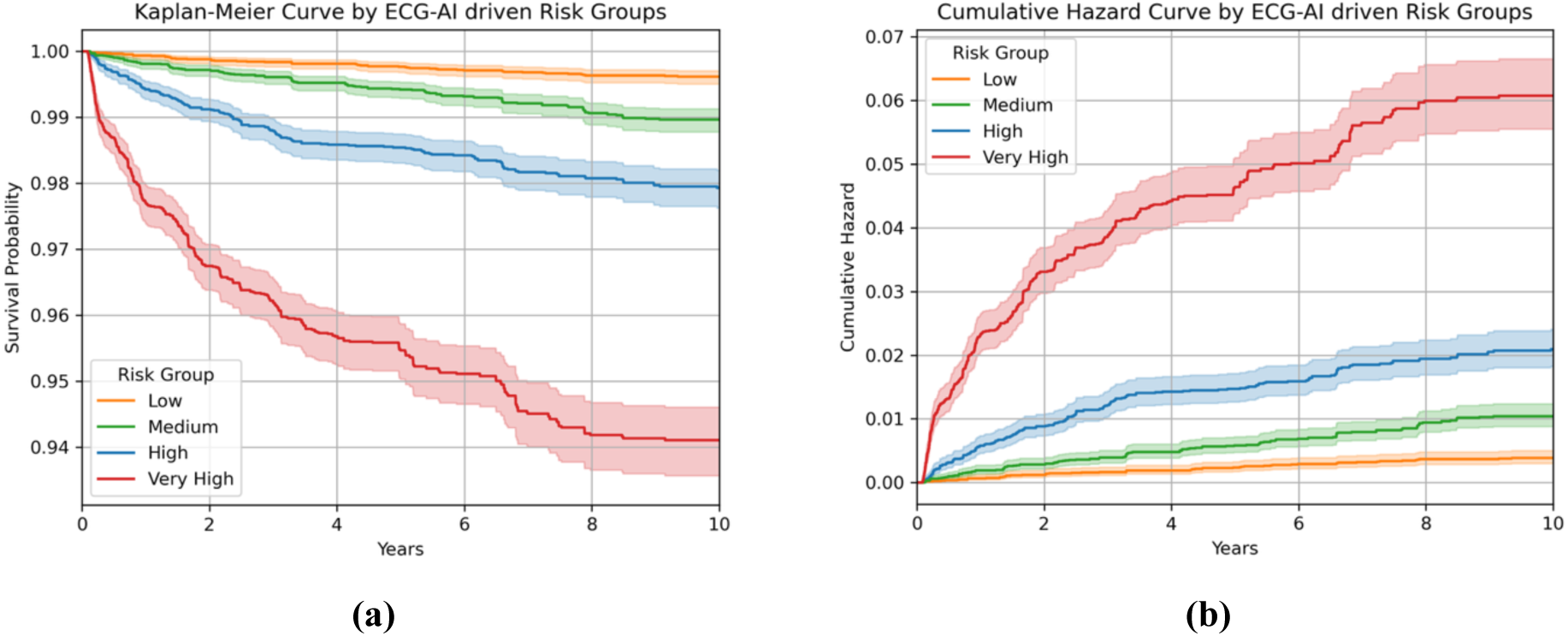
The Kaplan-Meier Survival Curve (a) and Nelson-Aalen Cumulative Hazard Curve (b) for the ECG-AI Risk Groups for Incident HFpEF.

Consistent with these survival patterns, the ECG-AI categorical risk variable was strongly associated with incident HFpEF in the Cox proportional hazards model. In the unadjusted model, the hazard ratio (HR) was 2.61 (95% CI: 2.43–2.80; p < 0.005), with a concordance index of 0.76. The proportional hazards assumption was tested using Schoenfeld residuals and showed no evidence of violation (p = 0.9047). After adjusting for age, sex, and race (Black or African American), the model yielded a slightly attenuated but still robust association, with an adjusted HR of 2.40 (95% CI: 2.23–2.58; p < 0.005) for the ECG-AI risk strata and an improved concordance of 0.79. These findings suggest that the ECG-AI risk score is a strong and independent predictor of HFpEF onset, even after accounting for key demographic factors.

## DISCUSSION

### Main Findings

This multi-stage evaluation demonstrates that a ECG-AI model, trained solely on real-world EHR data, can (i) accurately identify HFpEF in an external data with rigorous HFpEF annotations (HeartShare AUC = 0.76), (ii) the single lead version of ECG-AI produce almost identical accuracy (AUC of 0.77) as a promise for remote applications via wearables (iii) produce highly reproducible risk estimates across sequential ECGs (Pearson ≈ 0.88; κ ≈ 0.71) suggesting within patient reproducibility, and (iv) signal HFpEF risk as early as four years before clinical diagnosis with no material loss of discrimination (AUCs ranging from 0.77 – 0.80). Survival analyses demonstrated a statistically significant association between ECG-AI predictions and risk for incident HFpEF. Notably, this association remained robust after adjustment for conventional risk factors, with hazard ratios of 2.61 and 2.40, respectively, indicating that the ECG-AI output functions as an independent predictor of HFpEF. Together, these results indicate that a low-cost, easily deployed single lead ECG tracing, which is already embedded in many consumer wearables, can deliver longitudinal insight into HFpEF risk long before conventional diagnostic pathways are triggered.

### Existing Literature

Most ECG-AI publications in the HF domain focus on reduced ejection fraction or treat HF as a composite outcome, typically require the full 12-lead recording, and rarely undergo rigorous external validation, especially utilizing NIH-funded cohort studies with rigorous outcome annotations (23,30,34–36). A few HFpEF-specific models have emerged (23,24) but are limited by small sample size, lack of rigorous adjudication, or absence of repeat-ECG analyses. Kwon et al. (25) leveraged echocardiographic parameters to define HFpEF and developed a 12-lead ECG-based predictive model, achieving an AUC of 0.87. However, the proximity of ECG and echocardiogram acquisitions (within 1 week) may not accurately represent real-world clinical scenarios, where temporal disparities between diagnostic tests are common. In contrast, Hong et al. (26) employed a broader time window of ±1 year for pairing 12-lead ECGs with echocardiograms, yielding an AUC of 0.81. Notably, when ECGs recorded within 3 months of echocardiography were excluded, the AUC remained relatively stable at 0.80. Their HFpEF definition relied on the HFA-PEFF score (37), with cases defined as HFA-PEFF ≥ 5 and controls as HFA-PEFF < 5. However, the HFA-PEFF score has inherent limitations, including inadequate sensitivity, and its application may lead to diagnostic inaccuracies in detecting HFpEF, as previously reported (38–41). Our work closes each gap by leveraging (i) HeartShare’s rigorous HFpEF phenotyping, (ii) triplicate ECGs per participant to test reproducibility, and (iii) adaptability for both 12 lead and single lead (lead I) ECGs, thus aligning with single-lead wearables (e.g., Apple Watch) and adhesive patches within platforms such as ECG-Air (42).

### Clinical Interpretation of Error Analysis

Correctly classified HFpEF cases displayed faster atrial/ventricular rates, longer QRS and QTc intervals, and larger T-wave axes than misclassified cases. These patterns are consistent with heightened sympathetic tone, latent conduction slowing, and repolarization heterogeneity, as often observed in hypertensive or fibrotic ventricles—features that the neural network evidently exploits. Conversely, false-negative cases exhibited more “normal-like” ECGs and lower prevalence of hypertension, atrial fibrillation, and elevated filling pressures, suggesting that ECG-AI sensitivity wanes when electrical remodeling is minimal or absent. Integrating clinical variables or other physiological could bolster detection of these subtle phenotypes.

### Clinical Implications

HFpEF is under-detected in the community, and even among those identified, there are delays in diagnosis. Integration of such ECG-AI models with cardiology information system and/or wearables, when used together patient reported symptoms and other clinical data, may help with early recognition of HFpEF leading to timely interventions and improved outcomes. Such ECG-AI model can also help for more targeted recruitment for clinical trials of novel interventions to reduce transition from preclinical HFpEF to overt symptomatic HFpEF.

### Limitations

Our study has several limitations. First, the overall HFpEF detection accuracy is modest. Integration of patient reported symptoms and other clinical risk factors may lead to more clinically actionable precision. Second, while our results are all based on lead I ECG, which is mimicked by the smartwatches with ECG functionality, future studies are needed to assess concordance of clinical lead I and single lead wearable ECGs. Despite previous studies showed such concordance for other conditions such as fatal coronary heart disease, new studies specifically for HFpEF detection from wearables are needed. While the potential deployment of a single-lead model represents a promising step toward broader clinical translation, it is important to acknowledge that financial and technological barriers—such as access to wearable devices and the necessary digital infrastructure—may still limit equitable implementation, particularly in underserved populations. Third, medications, comorbidities, or device noise are not accounted for in this ECG-AI model, data could influence predictions. Fourth, cases and controls were not verified using the gold standard of invasive exercise testing, but HeartShare employed high specificity criteria including elevated H2FPEF (12) or HFA-PEFF scores (37), increasing confidence in the veracity of the HFpEF diagnosis. Lastly, the clinical interpretation of error analysis identified certain ECG characteristics significantly different between correctly and misclassified HFpEF cases. However, we note that such ECG differences are not necessarily only related to HFpEF. There is a need for such analysis where non-HFpEF cases include individuals with a broader range of cardiovascular diseases.

## CONCLUSIONS

ECG-AI model delivers robust, reproducible, and remarkably early detection of HFpEF across diverse populations from both 12 lead and single lead ECG. By transforming a ubiquitous, inexpensive signal into a powerful risk-stratification tool, this approach holds promise to shift HFpEF care from late, resource-intensive diagnosis toward early, preventive intervention. Prospective studies will determine whether the model chiefly unmasks undiagnosed HFpEF or forecasts future disease—and, ultimately, whether its integration into clinical practice improves patient trajectories and resource allocation.

## Data Availability

HeartShare data are part of an NIH-supported cohort study and are available to qualified investigators through the HeartShare data access mechanisms established by the NIH. Real-world ECG data from Wake Forest Baptist Health cannot be shared due to institutional policies and patient privacy protections. The models used in this study will be made available for reproducibility purposes upon reasonable request to the corresponding author.

